# Public Health Impact of the Pfizer-BioNTech COVID-19 vaccine (BNT162b2) in the first year of rollout in the United States

**DOI:** 10.1101/2022.02.24.22271478

**Authors:** Manuela Di Fusco, Kinga Marczell, Kristen A. Deger, Mary M. Moran, Timothy L. Wiemken, Alejandro Cane, Solène de Boisvilliers, Jingyan Yang, Shailja Vaghela, Julie Roiz

**Author notes:** **Corresponding author** Manuela Di Fusco, Health Economics and Outcomes Research, Pfizer, Inc. New York, NY, USA.

## Abstract

**Background:** As the body of evidence on COVID-19 and post-vaccination outcomes continues to expand, this analysis sought to evaluate the public health impact of the Pfizer-BioNTech COVID-19 Vaccine, BNT162b2, during the first year of its rollout in the US.

**Methods:** A combined Markov decision tree model compared clinical and economic outcomes of the Pfizer-BioNTech COVID-19 Vaccine (BNT162b2) versus no vaccination in individuals aged ≥12 years. Age-stratified epidemiological, clinical, economic, and humanistic parameters were derived from existing data and published literature. Scenario analysis explored the impact of using lower and upper bounds of parameters on the results. The health benefits were estimated as the number of COVID-19 symptomatic cases, hospitalizations and deaths averted, and Quality Adjusted Life Years (QALYs) saved. The economic benefits were estimated as the amount of healthcare and societal cost savings associated with the vaccine-preventable health outcomes.

**Results:** It was estimated that, in 2021, the Pfizer-BioNTech COVID-19 Vaccine (BNT162b2) contributed to averting almost 9 million symptomatic cases, close to 700,000 hospitalizations, and over 110,000 deaths, resulting in an estimated $30.4 billion direct healthcare cost savings, $43.7 billion indirect cost savings related to productivity loss, as well as discounted gains of 1.1 million QALYs. Scenario analyses showed that these results were robust; the use of alternative plausible ranges of parameters did not change the interpretation of the findings.

**Conclusions:** The Pfizer-BioNTech COVID-19 Vaccine (BNT162b2) contributed to generate substantial public health impact and vaccine-preventable cost savings in the first year of its rollout in the US. The vaccine was estimated to prevent millions of COVID-19 symptomatic cases and thousands of hospitalizations and deaths, and these averted outcomes translated into cost-savings in the billions of US dollars and thousands of QALYs saved. As only direct impacts of vaccination were considered, these estimates may be conservative.

**KEY SUMMARY POINTS:** *Why carry out this study?:*

- Assessing the population-level health and economic impact of the Pfizer-BioNTech COVID-19 Vaccine (BNT162b2) is important for policy makers and payers who support decision-making and investment in vaccination.
- These analyses may be relevant to the public, especially those who remain hesitant to COVID-19 vaccination.

*What was learned from the study?:*

- This study showed that Pfizer-BioNTech COVID-19 Vaccine (BNT162b2) was an effective strategy that contributed to generating substantial public health impact and economic gains in the US in 2021
- The vaccine was estimated to prevent millions of COVID-19 symptomatic cases and thousands of hospitalizations and deaths, and these averted outcomes translated into cost-savings in the billions of US dollars and millions of QALYs saved
- The study highlights the importance of continuing widespread uptake of the Pfizer-BioNTech COVID-19 Vaccine (BNT162b2) to prevent COVID-19 related disease and generate substantial benefits from a broad, patient-centric, societal perspective

## INTRODUCTION

The coronavirus disease 2019 (COVID-19) pandemic has claimed millions of lives globally [1], with the United States (US) sharing a significant burden of more than nine hundred thousand deaths during the past two years [2]. As a public health response to curb the pandemic, multiple mitigation strategies including mass vaccination have been implemented [3].

The clinical effectiveness of COVID-19 vaccines in preventing infections and severe disease has been established in randomized clinical trials (RCTs) [4-7], while a rich body of real-world evidence (RWE) complements RCTs results [8, 9]. Leveraging national surveillance data, studies quantified the population-level impact of COVID-19 vaccination in the US and showed that vaccines are clinically effective in reducing symptomatic cases, deaths, and hospitalizations [10-13]. However, only a few studies assessed the economic impact of COVID-19 vaccination in the US [14-19]. Kohli et al., [15] and Padula et al., [16], published in the early stages of the pandemic, assessed the cost-effectiveness of a hypothetical COVID-19 vaccine meeting the WHO and Food and Drug Administration (FDA) target product profiles. Li et al., [17] estimated the cost-effectiveness of a booster strategy of BNT162b2 in individuals aged ≥65 years. Although the three studies demonstrated cost-effectiveness versus no vaccination, they focused on the healthcare system perspective, which may arguably underestimate the full value of vaccination. COVID-19 causes productivity loss, with an estimated 10 sick days per infected person [20], and ∼5.4% reduction in the labor force participation in the US [21, 22]. Analyses under a societal perspective are recommended for a more comprehensive assessment, incorporating impact on patients and society as a whole [23, 24]. Bartsch et al., [18] estimated that a non-specific COVID-19 vaccine can save billions of US dollars in productivity losses. Incorporating the broader macro-economic impact (e.g., GDP loss, deferred care), Kirson et al., [19] estimated that COVID-19 vaccines can generate trillions in savings.

More than one-year through the US vaccination program, extensive RCTs and RWE data are available on the clinical profile of the three vaccines (Pfizer-BioNTech; Moderna; Johnson & Johnson/Janssen) authorized by the FDA [25-27]. Surveillance data has been widely available [28-30], and research on COVID-19 economic burden [31-34] has emerged. These studies can be holistically combined to generate more accurate estimates of the societal value of vaccination.

The Pfizer-BioNTech COVID-19 Vaccine (BNT162b2) was the first vaccine available under Emergency Use Authorization (EUA) since December 2020, and became licensed as a two-dose primary series in individuals aged ≥16 years in August 2021 [25, 35, 36]. It was the only vaccine with FDA approval in 2021 and the only vaccine authorized for children; in the 12-15 years age group since May 2021, and in the 5-11 age group since October 2021 [36]. Based upon the Centers for Disease Control and Prevention (CDC) estimates, six out of ten (∼57%) individuals fully vaccinated in the US received Pfizer-BioNTech COVID-19 Vaccine (BNT162b2) in 2021 [37]. Representing the highest proportion of the vaccinated population, its use and outcomes have been heavily investigated in large, and high-quality studies [37]. This comprehensive body of evidence enables further research into its societal impact.

We employed quantitative methods to assess the public health and economic impact of two-dose BNT162b2 in the first year of its rollout in the US. The main analysis focused on the population aged ≥12 years, as the 5-11 age group was not eligible during most of the study period.

## METHODS

### Study Design

A combined Markov and decision tree model with transition probabilities based on COVID-19 literature, was developed to estimate the one-year public health impact of Pfizer-BioNTech COVID-19 Vaccine (BNT162b2) compared to no vaccination. The base case analysis was conducted in the US target population aged ≥12 years, from a societal perspective. Age-stratified epidemiological, clinical, economic, and humanistic parameters were derived from published literature.

The expected public health impact was estimated as the number of COVID-19 deaths and symptomatic cases averted, with the latter differentiated among those requiring outpatient versus inpatient care. The expected economic impact was estimated as the amount of avoided healthcare and societal costs corresponding to the health outcomes averted. Productivity and QALY loss associated with early death was accounted for on a lifetime horizon.

The model was programmed in Microsoft Excel® 2021. The manuscript was developed in alignment with the Consolidated Health Economic Evaluation Reporting Standards (CHEERS) 2022 guidelines (Table S1) [38].

### Model Structure

A Markov model (Figure A) was constructed with eight mutually exclusive health states based on: i) Susceptible-Infected-Recovered (SIR) structure which has been used in monitoring and predicting the spread of COVID-19 [39], and ii) Vaccinated states of partial (dose 1) or full vaccination (dose 2) depending on the vaccine coverage and breakthrough infections. The ‘infected states’ (infected, infected post dose 1, infected post dose 2) of the Markov model were linked with a Decision tree (Figure B) for transition probabilities of symptomatic and asymptomatic infections, and sequelae such as hospitalization, receiving intensive care (ICU) and/or invasive mechanical ventilation (IMV), and post-acute sequelae of COVID-19 (PASC).

**Figure A:**
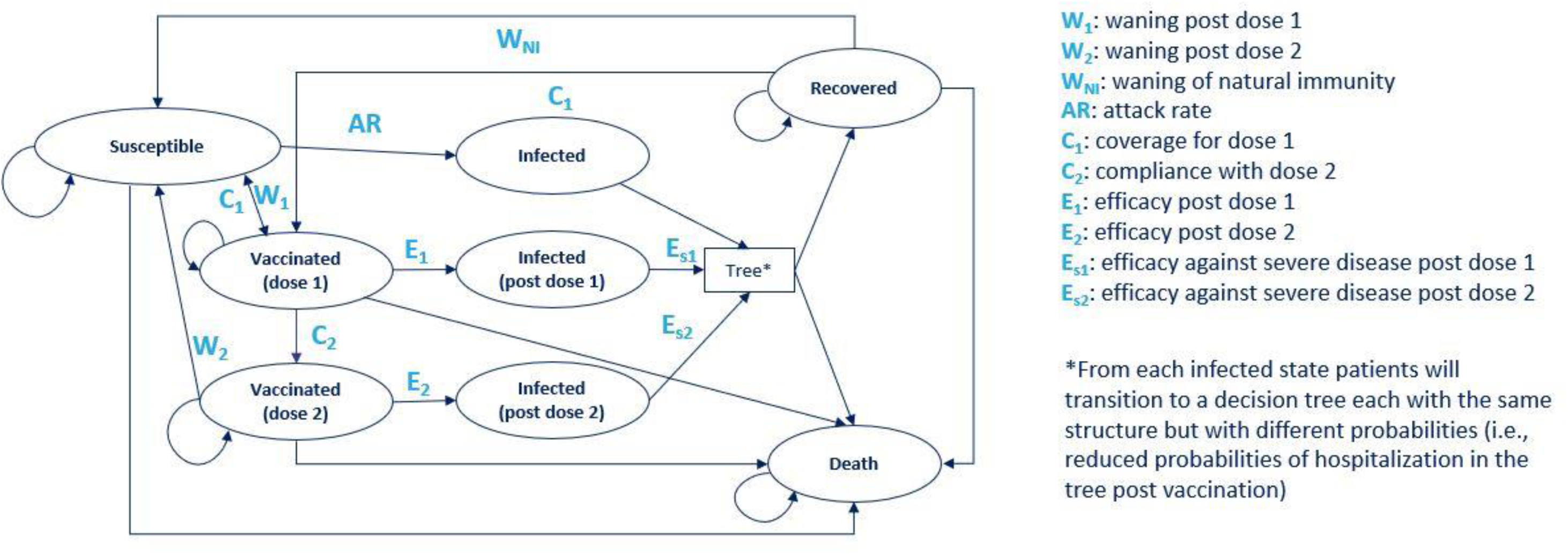
Structure of the Markov model.

**Figure B:**
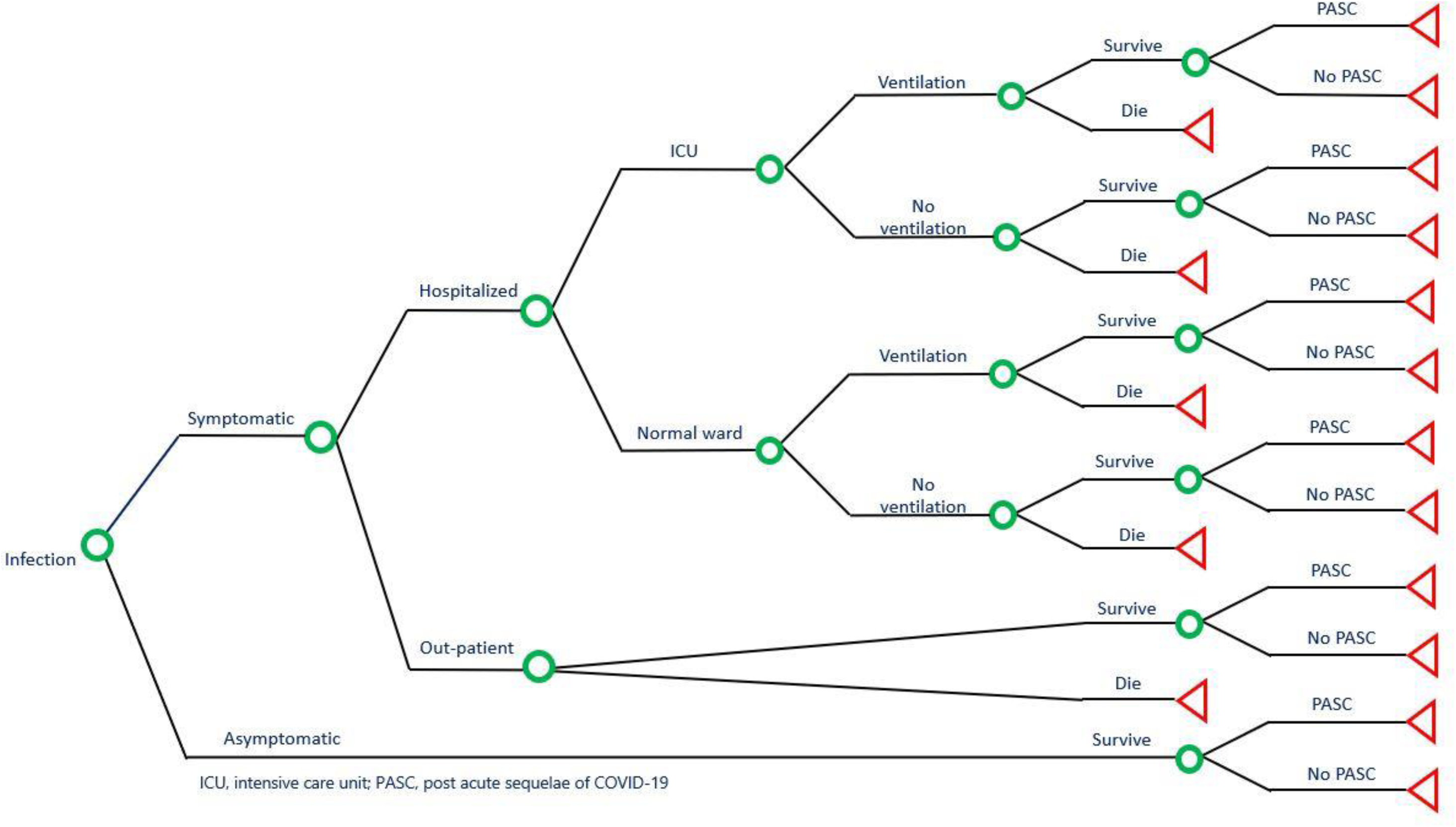
Decision tree for the probabilities of COVID-19 symptoms and sequelae.

Individuals entered the Markov model (Figure A; Table 1) in the ‘susceptible’ or ‘recovered’ health states with weekly cycles for one full year. At each weekly cycle, individuals could transition to other health states or remain in the current state. From the susceptible state, individuals could become infected or receive the vaccine (dose 1 with partial vaccination followed by dose 2 of full vaccination). An age-dependent yearly attack rate in the susceptible population defined the share of the model cohort that would become infected with COVID-19 and vaccination was determined by age-dependent vaccine coverage (further explained in model parameters). From the vaccinated states, patients could experience breakthrough infections or become susceptible again due to waning of vaccine protection. The former was governed by the vaccine efficacy, which could vary by dose, age group and the circulating variants of COVID-19. Waning of the efficacy of the vaccine and of infection-induced immunity was captured through their corresponding duration of protection. In each cycle of the Markov model, a fraction of the cohort was moved from the ‘vaccinated’ (1 or 2 doses) and ‘recovered’ health states to the ‘susceptible’ health state, representing the gradual decrease of protection level in the population due to waning.

**Table 1.**
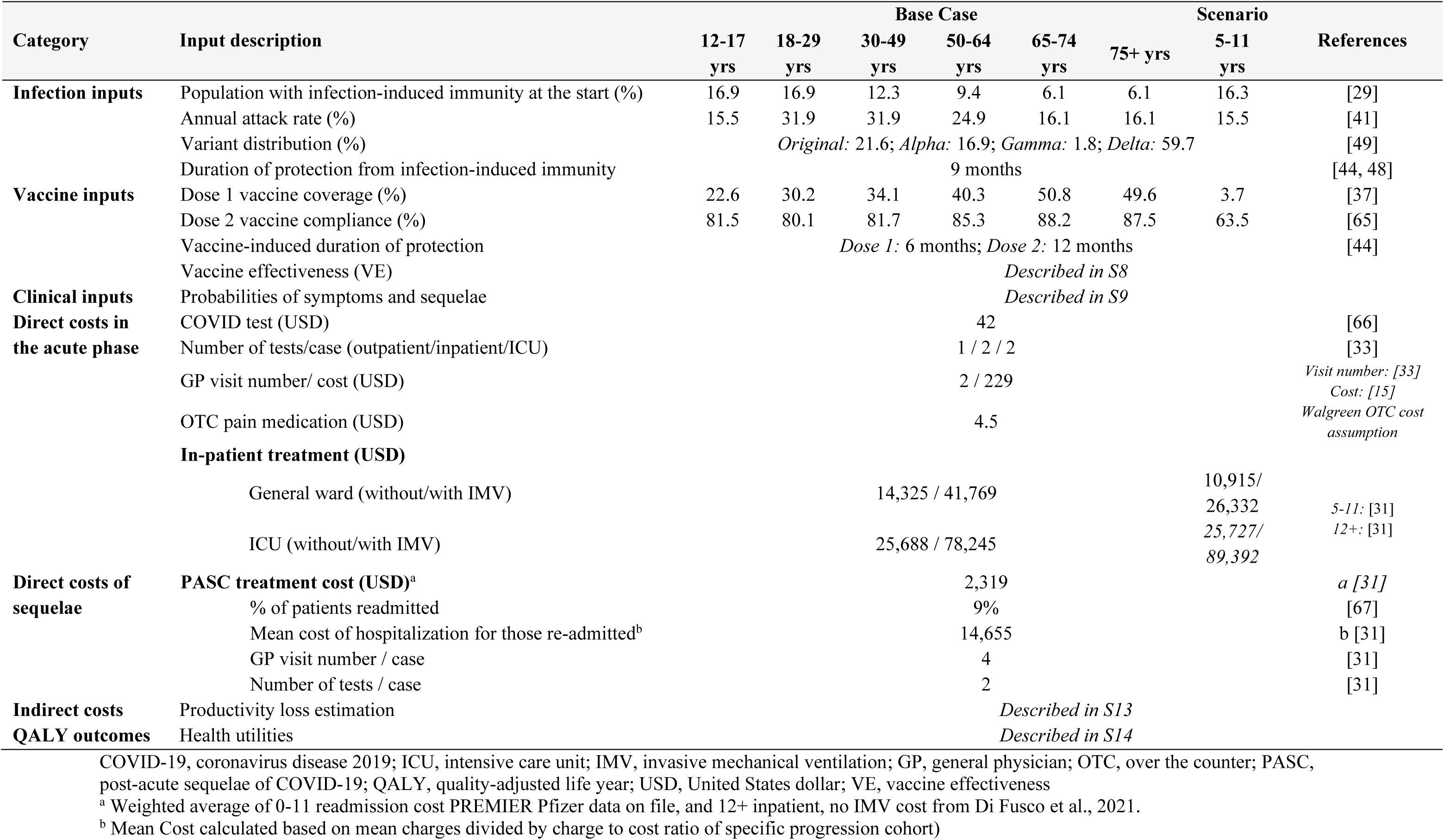
Model Parameters.

Once individuals got infected in ‘infected states’ (infected, infected post dose 1, infected post dose 2), they moved to the decision tree (Figure B; Table 1) for symptomatic or asymptomatic infection. We assumed that individuals with asymptomatic infection did not die or require any inpatient or outpatient treatment, but could experience PASC complications. The term PASC, also known as ‘long COVID’, is used to describe the post-acute sequelae and long-term symptoms that can be experienced from weeks to months by persons recovering from COVID-19 [40]. Symptomatic cases were moved further into the decision tree according to outcome probabilities informed by the individual’s health state specific efficacy against symptomatic disease and against hospitalization. The probability that infected individuals experienced symptoms was based on peer-reviewed literature [41]. Symptomatic cases were assumed to be managed in the outpatient or hospitalized setting, and incurred healthcare costs for clinical care including, respectively, visits, testing, medication, and hospitalization treatment. Hospitalized patients remained in the general ward or were admitted to the ICU and could receive IMV in either setting. Further, individuals that survived infection were then subject to a probability of developing PASC and incurred the costs of managing these symptoms. The PASC probabilities were sourced from published literature [42, 43]. We also assumed that individuals who got infected were immune to reinfection for an average of nine months after the infection [44].

At the end of the decision tree, individuals returned to the Markov model component to either the ‘recovered’ or ‘death’ state, based on the outcome of the decision tree. Death following infection in the decision-tree was due to COVID-19, which reflected the case fatality rates (Table 1, Table S9), while death from any other state was due to all other cause mortality. The ‘recovered state’ represented individuals that had recovered from infection and had infection-induced immunity to disease.

### Model Parameters

The model included a multitude of inputs derived from published literature and supported by medical review. The main inputs are reported in Table 1, further details are included in the electronic supplementary material (Tables S2-14).

#### Population Demographics

The study population was stratified into six age groups (12–17 years, 18–29 years, 30–49 years, 50–64 years, 65–74 years, and ≥75 years) to reflect age-dependent outcomes of COVID-19, as well as to facilitate age-dependent scenarios for vaccination coverage for dose 1, compliance with dose 2, and vaccine efficacy. Estimates of population sizes were adopted from the US Census Bureau population projections for 2019 [45] (Table S2).

#### Infection inputs

The model is based on an estimate of the proportions of susceptible and recovered individuals at the start of 2021, as well as on an estimate of the proportion of susceptible that would have been infected with SARS-CoV-2 over the year without vaccination.

The US infection- and vaccine-induced SARS-CoV-2 seroprevalence data from Jones et al., [46] and the Morbidity and Mortality Weekly Report (MMWR) [47] were used to define the proportions with infection-induced immunity at model start (Table 1, S3). The case projections were informed by estimates of the risk of COVID-19 outcomes before any COVID-19 vaccines became available; specifically, the probability of infection in the susceptible population was calculated from age-dependent one-year attack rates extrapolated from Reese et al., [41] (Table S4). Once the cases were distributed according to the demographic structure previously described, age-stratified data reported by the CDC during the vaccination era [41] was used to assess face validity and inform scenario analyses.

Similarly, in the absence of age-, variant- and dose-stratified data on the infection-induced duration of protection, it was assumed that duration of protection from infection-induced immunity would last for nine months. These assumptions were based upon emerging data on infection-induced long-term protection [44, 48] and medical opinion, and were tested in scenario analyses.

The CoVariants – Global initiative on sharing all influenza data (GISAID) database [49] was used to calculate the COVID-19 variant distribution. The frequencies of COVID-19 original strain, Alpha, Beta, Gamma, and Delta during different months were extracted to estimate their respective proportions over the year (Table 1, Table S6). The proportion of sequences for Beta was close to 0% hence this variant was not included due to its very low circulation in the US [49]. Additionally, data from the CDC was used to assess face validity [37].

#### Vaccine inputs

Using the two-dose series of the Pfizer-BioNTech COVID-19 Vaccine (BNT162b2) as the intervention, vaccine-specific inputs such as coverage, compliance with the second dose, VE, and duration of protection, were integrated from various sources and based upon medical opinion. Consistent with current vaccine implementation efforts, we assumed individuals were eligible for vaccination regardless of prior infection.

For estimating the age group-specific vaccination coverage rates of dose 1 and dose 2, the demographic trends of individuals aged >=12 years receiving COVID-19 vaccinations in the US reported by the CDC COVID-19 Data Tracker were used [50], wherein, coverage by age was averaged over 2021 to estimate the 12-month representative rates, using the area under the curve (AUC) method (Table S6). The age group-specific rates of vaccine coverage (defined as the percentage of eligible population receiving first dose of primary vaccination during model horizon) and vaccine compliance with second dose (defined as the percentage of population receiving second dose of primary vaccination during model horizon, among those receiving primary vaccination) were estimated from the CDC Data Tracker [50] (Table S7).

Vaccine effectiveness (VE) of dose 1 and dose 2 against infection, symptomatic disease, and hospitalization was sourced from different RWE observational studies from the US and, whenever needed, other countries (See supplement section 2.6 and Table S8 for further details). VE from RWE was chosen instead of vaccine efficacy from RCTs because RWE had the additional granularity required for the model projections (i.e., across study periods relevant to the multiple variants that were prevalent in 2021). Though the VE varied by COVID-19 variant and endpoint, it was assumed to be equal across age groups in the absence of combined age-, variant- and dose-stratified data (Table 1, Table S8). Similarly, in the absence of age-, variant- and dose-stratified data on the vaccine duration of protection, it was conservatively assumed that the dose 1 and the dose 2 would provide protection for, respectively, 6 and 12 months until complete loss of efficacy. These assumptions were based upon emerging data on vaccine-induced and infection-induced long-term protection [44, 48], and medical opinion, which were also tested in scenario analyses using the lower and higher estimates.

#### Clinical inputs

As individuals from the infected states moved to the decision tree, probabilities of experiencing symptoms and sequelae were derived from published sources (Table 1, Table S9). The probability of experiencing symptoms and the age-stratified probabilities of hospitalization among symptomatic individuals were derived from Reese et al., [41]. A retrospective analysis of the US Premier Healthcare database (COVID-19 Special Release) by Di Fusco et al., [31] was used for the distribution of symptomatic cases by general ward or ICU and receipt of IMV, case fatality rates, and costs and length of stay for hospitalization. The mortality rates in the outpatient care setting were sourced from a retrospective analysis of the US Optum database (Table S9) [33].

#### Healthcare resource utilization (HCRU) and cost inputs

We incorporated direct and indirect costs associated with the averted health outcomes (Table 1, Tables S11-13). Direct medical costs included COVID-19 testing, disease management in inpatient (hospitalization) or outpatient (general physician [GP] visits, emergency room, and over-the-counter medication) setting. Hospitalization costs, stratified by general ward or ICU, and receipt of IMV, were sourced from Di Fusco et al., [31]. The cost of managing PASC was also included as a one-off aggregated cost accrued at the time of infection, which included cost elements for number of GP visits, number of COVID-19 tests, the percentage of patients readmitted [67] and/or experiencing PASC. Due to uncertainty around the long-term outcomes, scenario analyses tested the lowest and highest reported re-admission rates (“Low”: 4%, “High”: 15%) [67].

Indirect costs included productivity loss due to short-term illness and premature death (Table S13). Lost productivity costs due to illness were calculated based on the workforce participation rate, labor cost per week, and number of working days lost. The indirect cost associated with PASC based on working time lost for PASC patients was also included in the societal perspective. Long-term costs were discounted at a rate of 3% per year in accordance with economic evaluation guidelines from the Advisory Committee on Immunization Practices (ACIP) [24] and the Second Panel on Cost-Effectiveness in Health and Medicine [23]. All costs were expressed as 2020 US dollars.

#### Health utilities

Quality-adjusted life years (QALYs) were estimated by applying utility decrements to the short and long-term symptomatic outcomes, and subtracting them from the general population age-dependent utility norms (Table S14). In the absence of robust COVID-19 specific disutility weights, we used proxies from other infectious diseases such as *Clostridium difficile* infection and influenza, which have been used in previously published COVID-19 studies [15, 17, 51]. Long-term QALY loss associated with early death was included on a lifetime horizon and discounted at 3% annually. Further details on health utilities are available in the supplement section 2.9.

### Analyses

The base case predicted the health outcomes of the Pfizer-BioNTech COVID-19 Vaccine (BNT162b2) in terms of the number of COVID-19 symptomatic cases, hospitalizations, and deaths averted over a year, alongside the corresponding direct healthcare and productivity costs saved, and QALYs gained, compared with no vaccination, from a societal perspective.

Two scenario analyses were performed to assess the impact of the parameter uncertainty and to evaluate the robustness of the base case findings. The scenarios used extreme but plausible ranges of parameters, which were informed by the lower and the upper bounds of confidence intervals (CIs) of the values used in the base case, or, in their absence, by medical opinion. The lower bounds were used for a “Low” scenario, and the upper bounds were used for a “High” scenario for all parameters except those related to the proportion of individuals with infection-induced immunity at model start, and the duration of protection from infection-induced immunity; for both the parameters, their upper bound values were used in the “Low” scenario. The parameters tested were those carrying higher uncertainty due to wider CIs, or limited evidence. These included the inputs determining the clinical course of COVID-19 in the absence of vaccination (i.e., attack rate) (Table S4), the proportion of subjects with infection-induced immunity (Table S3), the duration of infection-induced immunity (Table 1), the probability of symptoms (Table S9), the hospitalization rate for new admissions (Table S9) and re-admissions (Table 1), and the probability of outpatient deaths (Table S9). The inputs related to the clinical profile of the vaccine were varied as well: the variant and dose-specific VE against infection, VE against symptomatic disease, VE against severe disease (Table S8), as well as dose-specific duration of vaccine-induced protection (Table 1). Rapid literature review efforts highlighted that a rich body of evidence exists on the burden of disease and the clinical effectiveness of the vaccine, and data on healthcare resource utilization (HCRU) and costs is quickly emerging. As additional evidence was reviewed, alternative plausible ranges for model inputs were derived and used in sensitivity analyses to identify the most sensitive parameters and to characterize the uncertainty. As such, extensive sensitivity analyses were conducted on over 15 model parameters to independently evaluate the impact of individual parameters on the baseline results, using values from alternative sources. One or multiple parameters within the same category were changed at a time using different sets of input values (Tables 4, Tables S2-12).

On an exploratory basis, the sensitivity analyses also examined the impact of the year-end rollout in children aged 5-11, building on the growing clinical and economic evidence in this age group (Tables S2-14).

Implementation costs were also considered on an exploratory basis. This study focused on the assessment of the preventable costs associated with vaccine-preventable disease; it did not consider implementation costs, such as those related to infrastructure, supply, training and human resource management, outreach, or ancillary care management. These costs have been considered largely unknown, and not a primary driver for decision-making on the authorization of COVID-19 vaccines in the context of the pandemic [52]. The cost of vaccine administration for the US payer system has however been pointed out [52], and adverse events have been reported in the RCTs [53, 54]. Sensitivity analyses explored the effects of these two additional cost categories on the overall cost saving estimates (Tables 4, Table S12).

Finally, sensitivity analyses were run to assess the impact of alternative utility decrements and duration of symptoms input values on the QALY loss. Given that literature on COVID-19 specific utilities is scarce, arbitrary values (± 10%) informed the sensitivity analyses on utility decrements. A longer duration of symptoms was tested: 14 days [15], versus 5 days used in the base case (Table S14).

## RESULTS

### Burden of disease in the absence of vaccination

Table 2 shows the estimated burden of disease in the absence of COVID-19 vaccination in the base case, “Low” and “High” scenarios.

**Table 2.** Estimated burden of disease in the absence of COVID-19 vaccination.

| Outcomes | “Low” scenario | Base case | “High” scenario |
| --- | --- | --- | --- |
| **Symptomatic cases, n** | 42,171,670 | 59,507,414 | 89,332,715 |
| **Deaths, n** | 323,950 | 497,001 | 809,834 |
| **Outpatient cases, n** | 39,869,272 | 56,041,320 | 83,776,394 |
| **Hospitalizations, n** | 2,302,398 | 3,466,094 | 5,556,321 |
| General ward without IMV | 1,696,424 | 2,555,808 | 4,100,063 |
| General ward with IMV | 99,736 | 149,710 | 239,488 |
| ICU without IMV | 215,747 | 324,707 | 519,978 |
| ICU with IMV | 290,491 | 435,870 | 696,792 |
| **QALY lost, discounted** | 3,790,162 | 5,742,161 | 9,234,445 |
| **Cost outcomes, USD** |  |  |  |
| Direct costs (billions) | \$108.0 | \$180.9 | \$302.4 |
| Productivity loss (billions) | \$193.3 | \$278.9 | \$426.5 |

ICU, intensive care unit; IMV, invasive mechanical ventilation; LY, life year; QALY, quality-adjusted life year; USD, United States dollar

It was estimated that, by the beginning of the vaccination campaign, about 33 million (“Low”: 31 million, “High”: 35 million) US individuals aged ≥12 and older had been infected with SARS CoV-2 and recovered, representing ∼12% of the eligible population. This approximation was based on a seroprevalence study [31], and was conservative, although consistent with prior literature (11.9% by 30^th^ October according to Sullivan et al., [55], and 14% by mid-November according to Angulo et al., [56]).

Leveraging nationwide COVID-19 burden of disease data reported prior to the introduction of COVID-19 vaccination [31], the base case and scenarios estimated that, of the eligible and susceptible population, 24.2% (“Low”: 17%, “High”: 31.8%) would have become infected with COVID-19 during 2021 in the absence of vaccination. These estimates were consistent with CDC burden of disease data for 2021 including the vaccination period (21.7% based upon [2]) as well as seroprevalence data [57].

Based upon a review of genome sequencing data [49], it was calculated that about 22% of all infections were of the original strain, 17% were Alpha, ∼0% were Beta, 2% were Gamma, and almost 60% were Delta. This distribution was found to be consistent with additional literature [58, 59]. Using age-stratified probabilities of symptoms and hospitalization rates reported by Reese et al., [41] (Table 1), about 3.5 million (“Low”: 2.3 million, “High”: 5.6 million) hospitalizations were estimated to have occurred in 2021 in the absence of vaccination. This amount corresponds to a 5.8% (“Low”: 5.5%, “High”: 6.2%) overall symptomatic hospitalization rate across age groups, which aligns with prior literature (3.4% according to Angulo et al., [56]) and is consistent with a total of over 4.4 million hospitalizations since the start of the pandemic, per the CDC Tracker (accessed on 2/17/2022) [2].

Using age-stratified mortality rates reported in real-world retrospective studies [31, 33], we estimated that almost 500,000 (“Low”: 323,000, “High”: 810,000) deaths would have occurred in 2021 in the absence of vaccination. This estimate is consistent with a total of over 920,000 deaths since the start of the pandemic, per the CDC COVID Data Tracker (accessed on 2/17/2022) [2], and with published estimates of excess deaths among unvaccinated [60].

### Base Case results

Based upon the clinical and economic inputs set for the base case (Table 1), it was estimated that, compared to no vaccination, the two-dose series of the Pfizer-BioNTech COVID-19 Vaccine (BNT162b2) prevented almost 8.7 million symptomatic cases, averted approximately 690,000 hospitalizations, and saved over 110,000 lives (Table 3).

**Table 3.** Total cumulative number of health outcomes averted by BNT162b2, and associated cost-savings.

| Outcomes | “Low” scenario | Base case | “High” scenario |
| --- | --- | --- | --- |
| **Symptomatic cases, n** | 5,033,511 | 8,663,085 | 13,853,033 |
| **Deaths, n** | 45,760 | 111,380 | 202,797 |
| **Outpatient cases, n** | 4,771,447 | 7,973,287 | 12,590,046 |
| **Hospitalizations, n** | 262,065 | 689,797 | 1,262,987 |
| General ward without IMV | 189,680 | 501,191 | 918,794 |
| General ward with IMV | 12,401 | 32,104 | 58,512 |
| ICU without IMV | 24,876 | 65,279 | 119,376 |
| ICU with IMV | 35,108 | 91,223 | 166,305 |
| **QALY lost, discounted** | 489,548 | 1,144,091 | 2,056,257 |
| **Cost outcomes, USD** |  |  |  |
| Direct costs (billions) | \$12.7 | \$30.4 | \$57.4 |
| Productivity loss (billions) | \$22.4 | \$43.7 | \$73.3 |

ICU, intensive care unit; IMV, invasive mechanical ventilation; LY, life year; QALY, quality-adjusted life year; USD, United States dollar

Up to, respectively, 17.8%, 54.8% and 77.3% of the vaccine-prevented symptomatic cases, hospitalizations, and deaths were estimated to have occurred among individuals aged ≥65 years.

The total hospitalizations prevented were estimated to be composed mainly of stays in a normal ward without mechanical ventilation (501,191 [72.7%]), although the vaccine was also estimated to prevent more severe hospitalizations (32,104 [4.7%] stays in a normal ward with mechanical ventilation, 65,279 [9.5%] ICU stays without mechanical ventilation, and 91,223 [13.2%] ICU stays with mechanical ventilation). The reduction in COVID-19 burden led to an estimated total direct cost savings of $30.4 billion, driven by inpatient and PASC costs.

Overall, about 1.1 million discounted life-years (LY) and QALYs were gained. Around 85.5% of this QALY gain stemmed from avoiding COVID-19 mortality, while quality of life losses corresponding to morbidity from infection symptoms and long-term effects of mechanical ventilation had a moderate effect.

The observed reductions in COVID-19 symptomatic cases were also associated with productivity gains of $43.7 billion. Around 35.7% of this gain was related to the productivity loss due to early death and 64.3% to lost workdays during illness.

### Scenario Analyses

Table 3 shows the outcomes averted in the two alternative scenarios (“Low”, and “High”). Using the most conservative estimates of burden of disease and clinical effectiveness, the “Low” scenario estimated that the Pfizer-BioNTech COVID-19 Vaccine (BNT162b2) prevented over 5 million symptomatic cases, averted over 260,000 hospitalizations, and saved over 45,000 lives.

The “High” scenario estimated that the Pfizer-BioNTech COVID-19 Vaccine (BNT162b2) prevented over 13.8 million symptomatic cases, averted over 1.2 million hospitalizations, and saved over 200,000 lives.

### Sensitivity Analyses

Sensitivity analyses results are presented in Table 4. The model was most sensitive to attack rates and hospitalization rates, whereas, probability of symptoms, clinical effectiveness, HCRU, and cost had moderate impacts on the outcomes of interest.

**Table 4.**
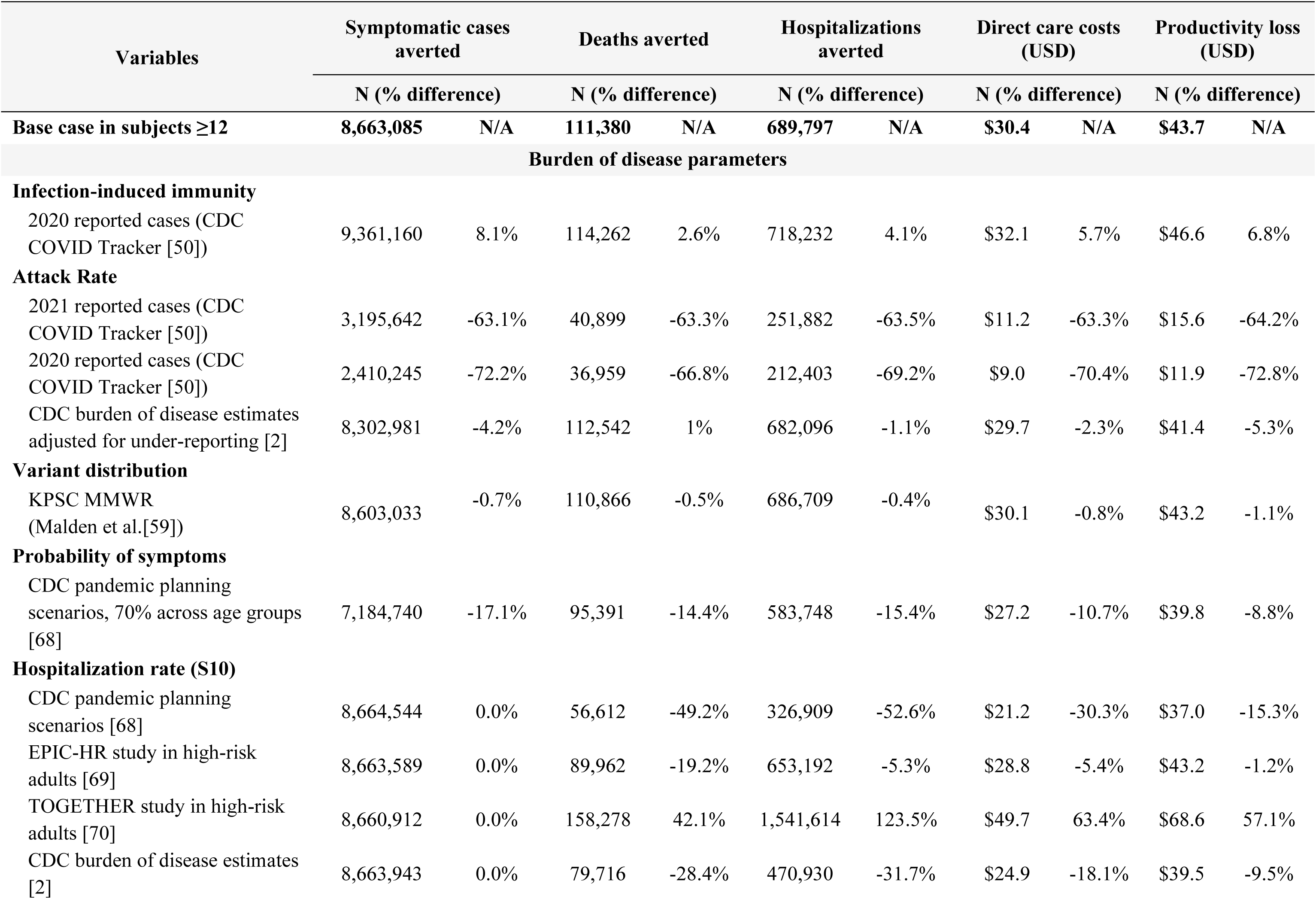
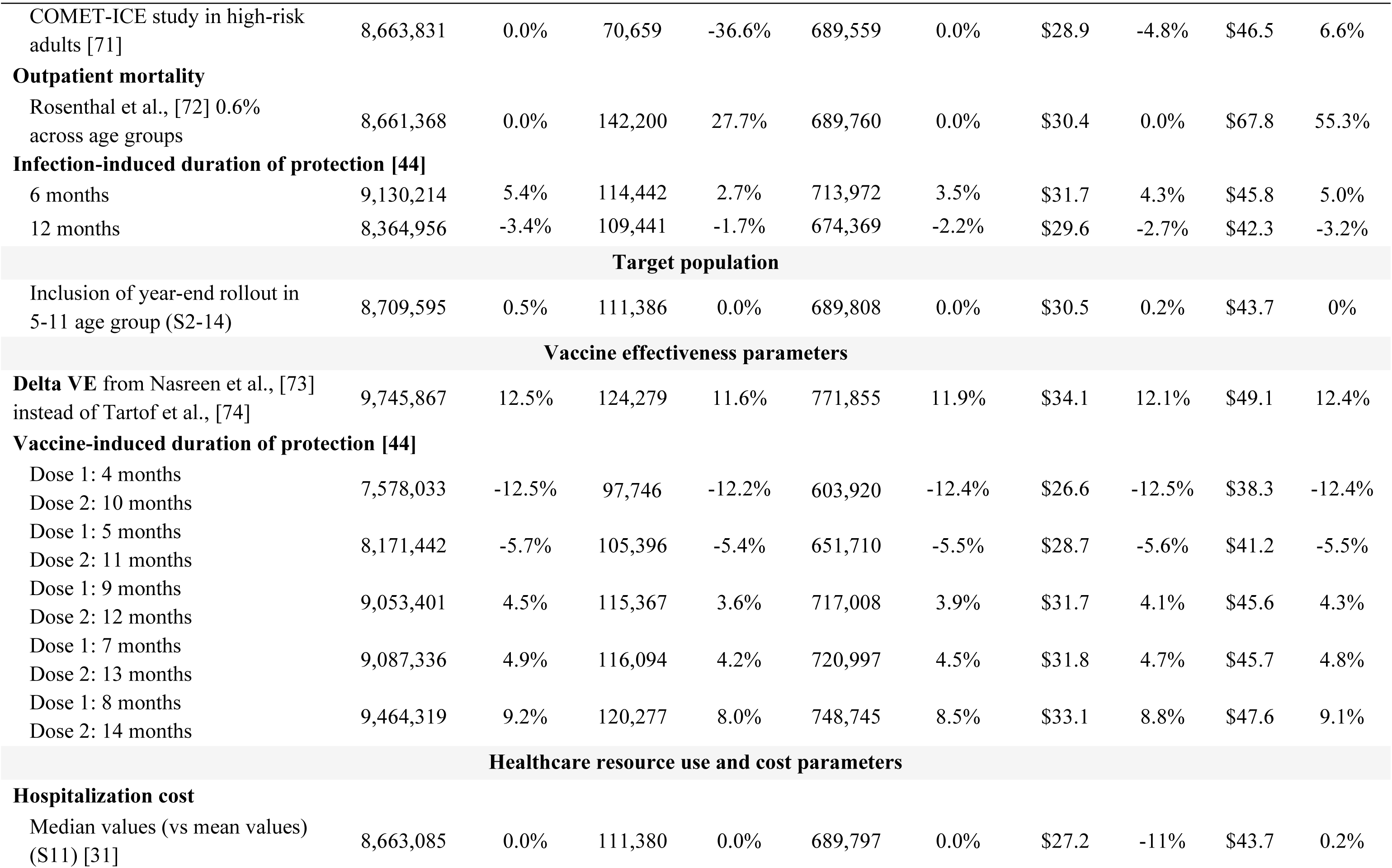
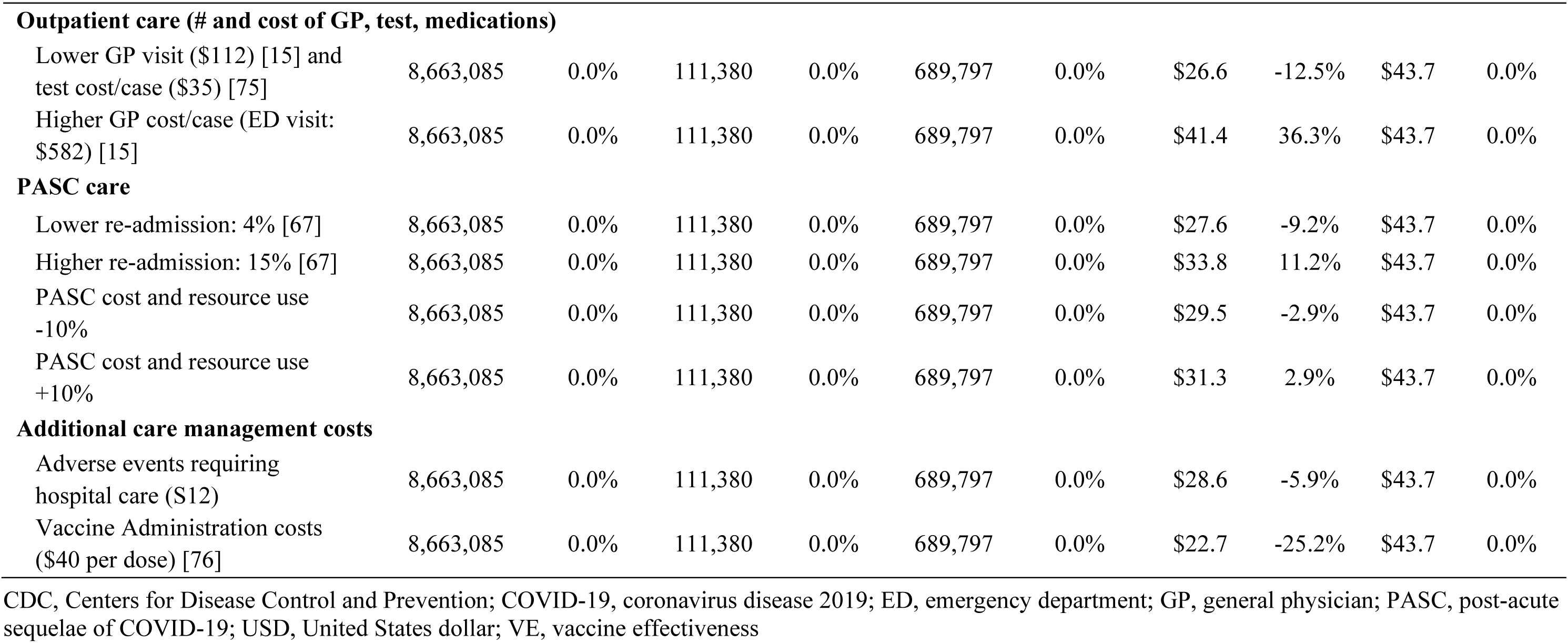
Sensitivity analysis.

As expected, attack rates defined based upon the CDC reported cases in 2020 or 2021 without adjusting for under-reporting led to significantly lower burden averted. The CDC burden of disease estimates adjusted for under-reporting were in line with the seroprevalence data used in the base case and had only a marginal impact. As under-detection and under-reporting especially in the early phase of pandemic have been widely documented [61], estimates of attack rates based on the reported cases are arguably not realistic and too conservative. As additional literature offered hospitalization rates across different time periods, age and risk-based populations, the hospitalization rates had a significant impact especially on the number of deaths and hospitalizations averted, which has led to a moderate to high effect on the direct costs and productivity loss. Alternative values for VE and vaccine-induced duration of protection moderately affected all the outcomes of interest, whereas, infection-induced duration of protection (i.e., +/- 3 months of base case at 9 months) had a marginal impact. The HCRU and costs had a moderate impact on direct care costs. Sensitivity analyses with 10% higher and lower utility decrement showed a marginal effect on the QALYs saved of ±0.2%. Increasing the duration of symptoms from 5 days (base case) to 14 days [15] led to a marginal increase of 3.2% in the QALYs saved.

The inclusion of the year-end rollout in children 5-11 slightly increased the outcomes averted, however, to a very low extent, driven by low vaccine coverage in the earliest phase of rollout in November and December 2021, as well as lower burden of disease than adults. Finally, the inclusion of additional care management costs (adverse events management and vaccine administration costs) had a low to moderate impact on the direct costs; those costs were offset by the total cost savings associated with vaccine-preventable disease.

## DISCUSSION

The analyses show that the Pfizer BioNTech COVID-19 Vaccine (BNT162b2) has had a profound public health impact in the US in 2021, by preventing millions of symptomatic cases, thousands of hospitalizations, and deaths, which translated into significant cost-savings in the billions of US dollars, considering direct costs only (i.e., payer perspective), as well as a societal perspective incorporative of productivity losses for the affected individuals. Such findings are consistent with other analyses that stressed the relevance of the social burden of disease when analyzing the outcomes [19]. This result was consistent in sensitivity and scenario analyses, and was highly impactful even excluding the potential indirect benefits of vaccination (linked to reduced transmission) [62], and the broader macroeconomic benefits associated with return to normalcy.

Quite similar to prior models [15-17], the results were most sensitive to inputs related to burden of disease and clinical effectiveness of the vaccine. Scenario analyses exploring parameter uncertainty showed that the base case results were robust. The most conservative estimates, represented in the “Low” scenario, confirmed the same trends and direction of results of the base case. The meaning and direction of the results also remained unchanged after running sensitivity analyses using alternative sets of input values. To our knowledge, this is the first study bringing together and simultaneously analyzing multiple types of outcomes and data (epidemiological, clinical, economic, and humanistic) to assess the public health impact of the Pfizer BioNTech COVID-19 Vaccine (BNT162b2) in the whole US eligible population. The study was informed by targeted literature review efforts that retrieved a large amount of recent and US-based data.

The quantitative analyses were based on a relatively simple, transparent, and interpretable model framework that was tailored to the purpose of the research question. The exploration of parameter uncertainty was conducted via extensive sensitivity analyses, whose results strengthened the findings of the base case and highlighted that, using either highly conservative or less conservative assumptions, the Pfizer-BioNTech COVID-19 Vaccine (BNT162b2) generated substantial health benefits and cost-savings.

The results of this study should, however, be considered in the context of several assumptions and limitations.

While the vast majority of the model inputs were derived from published COVID-19 literature and national surveillance data, uncertainty existed in parameter estimates and was tested in sensitivity analyses. Other forms of uncertainty (i.e., structural, and methodological) were not explicitly tested. The structures and mechanics used in the model were informed by medical opinion and were consistent with other published models [15-17], the existing data and understanding of the disease. Several simplifications and assumptions were made to minimize unnecessary complexity and deal with data limitations. Due to limited usable data on the spread of SARS-CoV-2 in the community and the effect of vaccination on transmission, our model simplified the epidemiology dynamics and time-dependent effects in a static Markov and decision tree model. Moreover, the model did not directly assess the transmission process and the indirect and herd immunity effects of vaccination, nor the potential therapeutic effects of vaccination in reducing the severity of cases and the impact of PASC, nor the potential macroeconomic savings that could have been achieved over the year through temporal elimination of levels of non-pharmaceutical interventions (NPIs) (e.g., restricted social interactions). Hence, this parsimonious model framework may have generated conservative estimates. These conservative estimates should, however, be considered in the context of a limited assessment of implementation costs, which was conducted on a small scale in a sensitivity analysis. Further studies assessing the implementation process and costs are warranted to improve the understanding of the full resource use associated with the COVID-19 vaccination program.

As this analysis used the Pfizer-BioNTech COVID-19 Vaccine (BNT162b2) as the intervention, results are not generalizable to other vaccines currently approved and authorized in the US, or in other countries. Similarly, the present study did not explicitly capture the effect of different existing or future interventions such as COVID-19 treatments, which may have a synergistic effect in further reducing disease severity.

Differently from existing studies [15-17], this study included variant-specific and outcome-specific VE for dose 1 and dose 2 of the Pfizer-BioNTech COVID-19 Vaccine (BNT162b2) (Table S7). VE estimates were partly informed by input data from other countries. Moreover, due to the absence of granular age-stratified data, VE estimates were assumed to be the same across the age groups, and no further stratification to risk groups was assessed. This is a limitation of the study, since RCTs and RWE show that VE varies across age and risk groups [63].

The vast majority of the inputs used were COVID-19 specific, however, in the absence of robust COVID-19 specific disutility weights, we used proxies from other infectious diseases such as *Clostridium difficile* infection and influenza, which have been used in previously published COVID-19 studies [15, 17, 51]. While this may increase uncertainty in the results, sensitivity analysis demonstrated that utilities had limited impact on the overall results.

The findings of this study may not fully reflect the current or future disease trends (e.g., prevalence of Omicron variant), vaccine clinical profile, healthcare resource use and costs. Results are neither generalizable to indications nor to populations those were not specifically covered in this analysis; those including children younger than 12, and specific high-risk populations such as immunocompromised who require additional vaccine doses for enhanced protection [64].

As COVID-19 data continues to rapidly expand and evolve, future studies are warranted to assess the public health impact in those additional settings as well as in the longer-term.

## CONCLUSIONS

This analysis shows that the Pfizer-BioNTech COVID-19 Vaccine (BNT162b2) generated substantial gains in health outcomes and cost savings in the US in 2021. It adds to the growing body of evidence demonstrating the societal benefit of the fast and extensive rollout of the vaccine in the US. It supports FDA and CDC recommendations for broad use of the vaccine, and highlights the opportunity to continue widespread uptake to prevent COVID-19 related disease and generate substantial benefits from a broad, patient-centric, societal perspective.

## Data Availability

Data generated or analyzed during this study are available upon request.

## Authorship

All named authors meet the International Committee of Medical Journal Editors (ICMJE) criteria for authorship for this article, take responsibility for the integrity of the work as a whole, and have given their approval for this version to be published.

## Authors’ Contributions

All authors contributed to the study conception, design, data acquisition, analysis, interpretation, and drafting and revising the manuscript.

## Medical writing, editorial, and other assistance

The authors acknowledge Moe Kyaw and Jessica Atwell (Pfizer Inc employees) for specific contributions to this research project. Editorial support was provided by Shailja Vaghela of HealthEcon Consulting, Inc and was funded by Pfizer.

## Compliance with Ethics Guidelines

These analyses used data from previously conducted studies and did not include data from any new studies with human participants or animals performed by any of the authors, hence ethical approval was not required.

## Availability of Data and Materials

Data generated or analyzed during this study are available upon request.

