## Supplemental material for "Public Health Impact of the Pfizer-BioNTech COVID-19 vaccine (BNT162b2) in the first year of rollout in the United States"

**Supplement material**

**Authors:** Manuela Di Fusco^1^, Kinga Marczell^2^, Kristen A. Deger^3^, Mary M. Moran^4^, Timothy L. Wiemken^4^, Alejandro Cane^1^, Solène de Boisvilliers^5^, Jingyan Yang^1,6^, Shailja Vaghela^7^, Julie Roiz^8^

**Affiliations:**

^1^ Pfizer Inc., New York, NY USA

^2^ Evidera, Bocskai út 134-146, Dorottya Udvar, E épület 2. emelet, Budapest, Hungary

^3^ Evidera, 7101 Wisconsin Ave., Suite 1400, Bethesda, USA

^4^ Pfizer, Inc, 500 Arcola Rd., Collegeville, PA, USA

^5^ Evidera PPD, 27 Rue Victor Hugo, 94200 Ivry-sur-Seine, France

^6^ Institute for Social and Economic Research and Policy, Columbia University, New York, NY, USA

^7^ HealthEcon Consulting, Inc., Ancaster, ON Canada

^8^ Evidera, 201 Talgarth Rd., Hammersmith, London, UK

**Corresponding author**

Manuela Di Fusco

Health Economics and Outcomes Research

Pfizer, Inc.

New York, NY, USA

ORCID ID: 0000-0003-0079-7331

1. **Consolidated Health Economic Evaluation Reporting Standards (CHEERS) Checklist 2022**

The Consolidated Health Economic Evaluation Reporting Standards (CHEERS) 2022 statement with a 28-item checklist [1], was used to prepare the manuscript. Each item reported in the main text or supplement is described below:

**Table S1: Consolidated Health Economic Evaluation Reporting Standards (CHEERS) Checklist 2022**

| **Section/topic** | **Item No** | **Guidance for reporting** | **Reported in section (pg no.)** |
| --- | --- | --- | --- |
| **Title** |  |  |  |
| Title | 1 | Identify the study as an economic evaluation and specify the interventions being compared. | 1 |
| **Abstract** |  |  |  |
| Abstract | 2 | Provide a structured summary that highlights context, key methods, results, and alternative analyses. | 2 |
| Introduction |  |  |  |
| Background and objectives | 3 | Give the context for the study, the study question, and its practical relevance for decision making in policy or practice. | 4-5 |
| **Methods** |  |  |  |
| Health economic analysis plan | 4 | Indicate whether a health economic analysis plan was developed and where available. | 5-6 |
| Study population | 5 | Describe characteristics of the study population (such as age range, demographics, socioeconomic, or clinical characteristics). | 5-8 |
| Setting and location | 6 | Provide relevant contextual information that may inﬂuence ﬁndings. | 5-11 |
| Comparators | 7 | Describe the interventions or strategies being compared and why chosen. | 5 |
| Perspective | 8 | State the perspective(s) adopted by the study and why chosen. | 5 |
| Time horizon | 9 | State the time horizon for the study and why appropriate. | 5 |
| Discount rate | 10 | Report the discount rate(s) and reason chosen. | 10-11 |
| Selection of outcomes | 11 | Describe what outcomes were used as the measure(s) of beneﬁt(s) and harm(s). | 5-11 |
| Measurement of outcomes | 12 | Describe how outcomes used to capture beneﬁt(s) and harm(s) were measured. | 5-11 |
| Valuation of outcomes | 13 | Describe the population and methods used to measure and value outcomes. | 5-11 |
| Measurement and valuation of resources and costs | 14 | Describe how costs were valued. | 10 |
| Currency, price date, and conversion | 15 | Report the dates of the estimated resource quantities and unit costs, plus the currency and year of conversion. | 10 |
| Rationale and description of model | 16 | If modelling is used, describe in detail and why used. Report if the model is publicly available and where it can be accessed. | 5-11, Supplement |
| Analytics and assumptions | 17 | Describe any methods for analysing or statistically transforming data, any extrapolation methods, and approaches for validating any model used. | 5-11, Supplement |
| Characterizing heterogeneity | 18 | Describe any methods used for estimating how the results of the study vary for subgroups. | 5-11, Supplement |
| Characterizing distributional effects | 19 | Describe how impacts are distributed across different individuals or adjustments made to reﬂect priority populations. | 5-11, Supplement |
| Characterizing uncertainty | 20 | Describe methods to characterize any sources of uncertainty in the analysis. | 11-12 |
| Approach to engagement with patients and others affected by the study | 21 | Describe any approaches to engage patients or service recipients, the general public, communities, or stakeholders (such as clinicians or payers) in the design of the study. | NA |
| **Results** |  |  |  |
| Study parameters | 22 | Report all analytic inputs (such as values, ranges, references) including uncertainty or distributional assumptions. | 5-12, Supplement |
| Summary of main | 23 | Report the mean values for the main categories of costs and results outcomes of interest and summarise them in the most appropriate overall measure. | 13-14 |
| Effect of Uncertainty | 24 | Describe how uncertainty about analytic judgments, inputs, or projections affect findings. Report the effect of choice of discount rate and time horizon, if applicable. | 15-16 |
| Effect of engagement with patients and others affected by the study | 25 | Report on any difference patient/service recipient, general public, community, or stakeholder involvement made to the approach or findings of the study | NA |
| **Discussion** |  |  |  |
| Study findings, limitations, generalizability, and current knowledge | 26 | Report key findings, limitations, ethical or equity considerations not captured, and how these could affect patients, policy, or practice. | 16-18 |
| **Other relevant information** | | | |
| Source of funding | 27 | Describe how the study was funded and any role of the funder in the identiﬁcation, design, conduct, and reporting of the analysis | 19 |
| Conﬂicts of interest | 28 | Report authors conﬂicts of interest according to journal or International Committee of Medical Journal Editors requirements. | 19 |

2. **Model parameters**
   2.1. **Population demographics**

**Table S2: Age-specific population**

| **Age Group**  **(years)** | **Population Size** | **Source** |
| --- | --- | --- |
| **5-11** | 28,384,878 | *US Census Bureau 2019* [2] |
| **12-17** | 25,135,943 |  |
| **18-29** | 53,728,222 |  |
| **30-49** | 84,488,200 |  |
| **50-64** | 62,925,688 |  |
| **65-74** | 31,483,433 |  |
| **≥75** | 22,574,830 |  |

US, Unites States

2.2. **Infection-induced immunity**

**Table S3: Infection-induced immunity**

| **Age group** | **Base Case (%)** | **Scenario Analysis (CI)^a^** | **Source** | **Sensitivity Analysis** | **Source** |
| --- | --- | --- | --- | --- | --- |
| **5-11^b^** | 16.3 | 12.9-22.0 | *Based on the estimated US Infection- and Vaccine-Induced SARS-CoV-2 Seroprevalence Data* [3] |  |  |
| **12-17** | 16.9 | 16.0-17.8 |  | 4.0 |  |
| **18-29** | 16.9 | 16.0-17.8 |  | 7.3 |  |
| **30-49** | 12.3 | 11.8-12.9 |  | 6.4 | *CDC COVID Data Tracker 2020 reported cases* [4] |
| **50-64** | 9.4 | 9.0-9.8 |  | 5.7 |  |
| **65-74** | 6.1 | 5.6-6.6 |  | 4.2 |  |
| **75+** | 6.1 | 5.6-6.6 |  | 5.1 |  |
| **Duration of protection of infection-induced immunity^c^** | 9 months | 8-10 months | [5, 6] | 6 months and  12 months | *Assumption based on the CDC science brief* [5] |

CDC, Centers for Disease Control and Prevention; CI, confidence interval; SARS-CoV-2, severe acute respiratory syndrome coronavirus 2; US, United States

^a^ ‘Low’ and ‘High’ scenario using the lower and upper bounds of CI, respectively.

^b^ Scenario analysis in the age group of 5-11.

^c^ Low and high scenario using the duration of protection due to natural immunity at 10 months and 8 months, respectively.

2.3. **Annual attack rates**

**Table S4: Annual attack rates used in the base case and “Low”/ “High” scenario analysis**

| **Age group** | **Annual Attack Rates, extrapolated from Reese et al.,[7]** | | | |
| --- | --- | --- | --- | --- |
|  | **Age group used in the source** | **Estimated Total Infections (rate/100,000)^a^** | **Extrapolated Attack Rates Base Case**  **%** | **Scenario Analysis**  **(CI)^b^** |
| **5-11^c^** | 5-17 | 10,336 | 15.5 | 11.8 - 20.9 |
| **12-17** | 5-17 | 10,336 | 15.5 | 11.8 - 20.9 |
| **18-29** | 18-49 | 21,292 | 31.9 | 23.7 - 44.9 |
| **30-49** | 18-49 | 21,292 | 31.9 | 23.7 - 44.9 |
| **50-64** | 50-64 | 16,601 | 24.9 | 18.7 - 34.1 |
| **65-74** | >65 | 10,750 | 16.1 | 11.8 - 23.0 |
| **75+** | >65 | 10,750 | 16.1 | 11.8 - 23.0 |

CI, confidence interval; SA, sensitivity analysis

^a^ Estimated infections per 100,000 reported in Reese et al. were for 8 months, which were adjusted to 12-month attack rates in the base case analysis.

^b^ ‘Low’ and ‘High’ scenario using the lower and upper bounds of CI, respectively.

^c^ Scenario analysis in the age group of 5-11.

**Table S5: Annual attack rates used in the sensitivity analysis**

| **Age group** | **Sensitivity analysis based on**  **the CDC 2020 cases [4]** | | **Sensitivity analysis based on**  **the CDC 2021 cases [4]** | | **Sensitivity analysis based on the CDC estimated COVID-19 burden [8]** |
| --- | --- | --- | --- | --- | --- |
|  | **Estimated symptomatic cases (rate/100,000)** | **Extrapolated 1-year attack rate (%)** | **Estimated symptomatic cases (rate/100,000)** | **Extrapolated 1-year attack rate (%)** |  |
| **12-17** | 3,999 | 4.9 | 10,432 | 10.0 | 19.1 |
| **18-29** | 7,307 | 8.9 | 11,972 | 12.0 | 29.5 |
| **30-49** | 6,424 | 7.8 | 11,331 | 11.0 | 29.5 |
| **50-64** | 5,660 | 6.9 | 8,594 | 9.0 | 23.5 |
| **65-74** | 4,236 | 5.1 | 6,345 | 6.0 | 16.6 |
| **75+** | 5,112 | 6.2 | 6,015 | 6.0 | 16.6 |

CDC, Centers for Disease Control and Prevention; COVID-19, coronavirus disease 2019

2.4. **Variant distribution**

**Table S6: Distribution of COVID variants based on the GISAID data**

| **Variant** | **Base Case** | | | **Sensitivity Analysis^a^** | |
| --- | --- | --- | --- | --- | --- |
|  | **Distribution %** | **Source** | **Distribution %** | | **Source** |
| **Alpha** | 16.9 | *CoVariants Database* [9] | 32.4 | | *KPMC MMWR*  *[10]* |
| **Gamma** | 1.8 |  | 7.2 | |  |
| **Delta** | 59.7 |  | 60.4 | |  |
| **Original Strain** | 21.6 |  | *NR* | |  |

CDC, Centers for Disease Control and Prevention; COVID, corona virus disease; GISAID, Global initiative on sharing all influenza data; KPMC, Kaiser Permanente Southern California; MMWR, CDC's Morbidity and Mortality Weekly Report; NR, not reported

^a^ Variant distribution was derived from the KPMC MMWR report during March 4-July 2021, while assuming same respective distribution Alpha/Gamma in Jan-Feb as March to July, and Delta emergence in March and dominance from August onwards. Data on original strain was not reported in the MMWR report.

2.5. **Vaccination coverage**

**Table S7: Age-specific vaccination coverage of BNT162b2**

| **Age group** | **BNT162b2 coverage dose 1 (%)** | **BNT162b2 coverage compliance with**  **dose 2 (%)** |  | **Source** |
| --- | --- | --- | --- | --- |
| **5-11^a^** | 3.7 | 63.5 |  |  |
| **12-17** | 22.6 | 81.5 |  | *CDC COVID Data Tracker*  [11] |
| **18-29** | 30.2 | 80.1 |  |  |
| **30-49** | 34.1 | 81.7 |  |  |
| **50-64** | 40.3 | 85.3 |  |  |
| **65-74** | 50.8 | 88.2 |  |  |
| **75+** | 49.6 | 87.5 |  |  |

CDC, Centers for Disease Control and Prevention; COVID, corona virus disease

^a^ Scenario analysis in the age group of 5-11.

2.6. **Vaccine effectiveness (VE)**

The transition rate from ‘susceptible’ and ‘recovered’ health states to ‘vaccinated’ (dose 1) health state was informed by the assumed vaccine coverage stratified by age group. This coverage, labeled as primary coverage defined the share of the eligible population receiving the first dose of primary vaccination during the model horizon. Compliance with the second dose indicated the share of the population receiving the second dose of primary vaccination during the model horizon, among those who received the first dose of primary vaccination during the model horizon or who entered the model already having received a first dose. The share of the population in the vaccinated (dose 1) health state at the start of the model was defined by the distribution of the starting population (see section Population). The primary vaccination was given at the start of the first year of the model.

As shown in the Table S7 below, Vaccine efficacy (VE) was adopted from different real world observational studies from the US and other countries. In the absence of data granularity, we estimated VE %(CI) for dose 1 and dose 2 for all the variants of interest, however, it was assumed to be equal across the age groups. Additional scenario analysis was performed using the confidence interval values to assess the impact of VE on the overall public health and economic value of BNT162b2 vaccine.

Vaccination affects the probability of infection, probability of symptomatic disease, and probability of hospitalization among infected, the latter representing efficacy against severe disease. Inputs provided for these three efficacy categories must increase in that order: efficacy against symptomatic disease must not be lower than efficacy against infection, and efficacy against hospitalization must not be lower than efficacy against symptomatic disease. No additional efficacy was calculated in the model for patients who had natural immunity and protection conferred by the vaccine was assumed to replace natural immunity.

Waning of the efficacy of the vaccine and of natural immunity was captured through the duration of protection. In each cycle of the Markov model, a fraction of the cohort was moved from the ‘vaccinated’ (1 or 2 doses) and ‘recovered’ health states to the ‘susceptible’ health state, representing the gradual decrease of protection level in the population due to waning. The corresponding transition rates were informed by the duration of protection. The duration of protection for dose 1 and dose 2 of all variants was assumed to be of 6 months and 12 months, respectively, based upon the emerging data on vaccine-induced and infection-induced long-term protection [5, 6]. Scenario analysis was performed using 10 months in the low scenario and 12 months in the high scenario for the one-year model horizon.

**Table S8: BNT162b2 Vaccine effectiveness based on RWE studies**

|  | **Vaccine Effectiveness based on the RWE studies for the Base Case (Scenario)^a^: %(CI)** | | | | | | | | | | | | | | | | |
| --- | --- | --- | --- | --- | --- | --- | --- | --- | --- | --- | --- | --- | --- | --- | --- | --- | --- |
| **Variant** | **Infection** | | | | **Symptomatic disease** | | | | **Hospitalization** | | | | **Death** | | | | |
|  | **1 dose** | **Ref.** | **2 dose** | **Ref.** | **1 dose** | **Ref.** | **2 dose** | **Ref.** | **1 dose** | **Ref.** | **2 dose** | **Ref.** | **1 dose** | **Ref.** | **2 dose** | **Ref.** | |
| **Original^b^** | 61.0  (50.8-69.2) | [12] | 88.0  (84.2-91.0) | [12, 13] | 67.0  (65.0-68.0) | [14] | 88.0  (84.2-91.0) | [12, 13] | 82.0  (81.0-84.0) | [14] | 96.0  (94.0-97.0) | [14] | 82.0  (81.0-84.0) | [14] | 96.0  (94.0-97.0) | [14] | |
| **Alpha** | 61.0  (50.8-69.2) | [12] | 88.0  (84.2-91.0) | [12, 13] | 67.0  (65.0-68.0) | [14] | 88.0  (86.0-90.0) | [14] | 82.0  (81.0-84.0) | [14] | 96.0  (94.0-97.0) | [14] | 82.0  (81.0-84.0) | [14] | 96.0  (94.0-97.0) | | [14] |
| **Delta** | 74.0  (55.0-85.0) | [15] | 75.0  (71.0-78.0) | [15] | 74.0  (55.0- 85.0) | [15] | 75.0  (71.0-78.0) | [15] | 79  (−49.0-97.0) | [15] | 93.0  (84.0-96.0) | [15] | 79^e^  (−49.0-97.0) | [15] | 93.0^e^  (84.0-96.0) | | [15] |
| **Gamma^c^** | 18.9  (−1.8-35.4) | [16, 17] | 74.3  (70.3-77.7) | [16, 17] | 63.0  (54.0-70.0) | [14] | 86.0  (0.0-98.0) | [14] | 80.0  (70.0-87.0) | [14] | 94.0  (59.0-99.0) | [14] | 80.0  (70.0-87.0) | [14] | 94.0  (59.0-99.0) | | [14] |
| **Delta - SA^d^** | 57.0  (53.0, 61.0) | [14] | 92.0  (89.0, 94.0) | [14] | 57.0  (53.0, 61.0) | [14] | 92.0  (89.0, 94.0) | [14] | 81.0  (76.0, 85.0) | [14] | 98.0  (96.0, 99.0) | [14] | 81.0^e^  (76.0, 85.0) | [14] | 98.0^e^  (96.0, 99.0) | | [14] |

CI, confidence interval; COVID-19, coronavirus disease 2019; RWE, real world evidence; SA, sensitivity analysis; VE, vaccine effectiveness

^a^ ‘Low’ and ‘High’ scenario using the lower and upper bounds of CI, respectively.

^b^ VE rates of Alpha variant were adopted for original variant in the absence of data.

^c^ VE rates against infection for Beta variant were adopted for Gamma variant in the absence of data.

^d^ VE rates for Delta variant in the sensitivity analysis were sourced from Nasreen et al., [14] instead of Tartof et al., [15] used in the base case analysis.

^e^ VE rates for hospitalization were adopted for deaths in the absence of data, as most studies [14, 16, 17] reported similar VE rates for hospitalization or death.

2.7. **Clinical inputs**

**Table S9: Probabilities of symptoms and sequelae**

| **Category** | **Input description** | **Base Case (%)** | | | | | | | **Scenario (%)^a^** | |
| --- | --- | --- | --- | --- | --- | --- | --- | --- | --- | --- |
|  |  | **12-17 yrs** | **18-29 yrs** | **30-49 yrs** | **50-64 yrs** | **65-74 yrs** | **75+ yrs** | **Ref.** | **5-11**  **yrs** | **Source** |
| **Clinical Inputs** | Probability of symptomatic infection | 85.2 | 85.3 | 85.3 | 85.1 | 80.8 | 80.8 | [7] | 85.2 | [18]  [7] |
|  | Hospitalization rate among symptomatic patients | 0.9 | 2.6 | 2.6 | 7.2 | 22.4 | 22.4 |  | 0.02 |  |
|  | **Among hospitalized patients:** | | | | | | | | |  |
|  | Critical care/ICU admission rate | 23.2 | 11.9 | 18.0 | 24.3 | 26.9 | 20.5 | [19] | 23.2 | [18] |
|  | IMV rate in General ward | 0.9 | 1.4 | 3.2 | 5.9 | 7.8 | 7.1 |  | 0.9 | [19] |
|  | IMV rate in ICU | 27.6 | 39.1 | 50.0 | 60.5 | 63.6 | 53.1 |  | 9.8 | [18] |
|  | **Probability of death among patients in:** | | | | | | | | |  |
|  | General ward without IMV | 0.13 | 0.02 | 0.15 | 1.13 | 4.25 | 14.14 | [19] | 0.10 | [19] |
|  | General ward with IMV | 0.0 | 15.1 | 25.1 | 40.2 | 53.5 | 66.7 |  | 0.0 |  |
|  | ICU without IMV | 0.0 | 0.6 | 0.8 | 3.6 | 10.4 | 26.1 |  | 0.0 |  |
|  | ICU with IMV | 9.1 | 20.0 | 33.9 | 48.2 | 60.4 | 69.4 |  | 1.8 | [18] |
|  | Outpatient care setting | 0.01 | 0.01 | 0.03 | 0.15 | 0.54 | 1.81 | [20]  *12-17: assumed the same as of age 18-29* | 0.01 | *assumed the same as of age 18-29* |
|  | **Probability of PASC among patients:** | | | | | | | | |  |
|  | Asymptomatic patients | 36.9 *(assumed the same rate for all age groups)* | | | | | | [21] | 36.9 | *Assumed the same as of adults* |
|  | Received outpatient care | 36.9 *(assumed the same rate for all age groups)* | | | | | | [21] | 36.9 |  |
|  | Received inpatient care | 44.8 | 45.7 | | | | | *12-17:* [22]  *18+:* [21] | 44.2 | [22] |

COVID19, coronavirus disease 2019; ICU, intensive care unit; IMV, invasive mechanical ventilation; PASC, post-acute sequelae of COVID-19

^a^ Scenario analysis in the age group of 5-11.

**Table S10: Hospitalization rates used in the sensitivity analysis**

| **Age group** | **Probability of hospitalisation among symptomatic patients (%)[23]** | **EPIC-HR (Paxlovid)**  **(High risk adults)**  **(%) [24]** | **MOVe-OUT (Molnupiravir)**  **(High risk adults)**  **(%) [25]** | **TOGETHER (Fluvoxamine)**  **(High risk adults)**  **(%) [26]** | **COMET-ICE (Sotrovimab)**  **(High risk adults)**  **(%) [27]** | **CDC Burden of disease estimates- Probability of hospitalisation among symptomatic patients (%) [8]** |
| --- | --- | --- | --- | --- | --- | --- |
| **5-11** | 1.7 | *N/A* | *N/A* | *N/A* | *N/A* | *N/A* |
| **12-17** | 1.7 | *NA* | *NA* | *NA* | *NA* | 0.76 |
| **18-29** | 1.7 | 4.8 | 1.5 | 11.7 | 7.2 | 1.97 |
| **30-49** | 1.7 |  |  |  |  | 1.97 |
| **50-64** | 4.5 |  |  | 21.9 |  | 5.43 |
| **65-74** | 7.4 | 16.3 | 12.6 |  |  | 13.96 |
| **75+** | 7.4 |  |  |  |  | 13.96 |

CDC, Centers for Disease Control and Prevention; COVID, corona virus disease; N/A, not applicable; NR, not reported

2.8. **Healthcare resource utilization and costs**

**Table S11: Inpatient/Hospitalization costs**

| **COVID-19 related Hospitalization cost** | **Base Case**  **(Mean USD)** | **Sensitivity Analysis (Median USD)** | **Source** |
| --- | --- | --- | --- |
| General ward without IMV | 14,325 | 9,504 | Di Fusco et al.,  [19] |
| General ward with IMV | 41,769 | 25,068 |  |
| ICU without IMV | 25,688 | 18,434 |  |
| ICU with IMV | 78,245 | 54,402 |  |

COVID19, coronavirus disease 2019; ICU, intensive care unit; IMV, invasive mechanical ventilation; -USD, United States dollars

**Table S12: Adverse Event (AE) rates and associated management costs used in the sensitivity analysis**

| **Adverse Event (AE)** | **Pfizer BioNTech (BNT162b2)** | | **AE management cost** | |
| --- | --- | --- | --- | --- |
|  | **AE Rates (%)** | **Source** | **(USD)** | **Source** |
| Myocarditis | 0.00094 | [28] | 32,295.0 | [29] |
| Pericarditis | 0.00240 | [28] | 14,465.0 | [29] |
| Myoperiocarditis | 0.00240 | *Assumed the same as of pericarditis* | 14,465.0 | *Assumed the same as of pericarditis* |
| Acute allergic reaction requiring hospitalization | 0.00055 | [30] | 7,727.6 | [29] |
| Disseminated intravascular coagulation | 0.0220 | [31] | 15,843.0 | [29] |
| Acute myocardial infarction | 0.0220 | [31] | 22,320.0 | [29] |

AE, Adverse event; USD, United States dollars

**Table S13: Indirect costs**

| **Category** | **Input description** | **Base case value** | | | | | | **Source** |
| --- | --- | --- | --- | --- | --- | --- | --- | --- |
|  |  | **5-17 yrs^a^** | **18-29 yrs** | **30-49 yrs** | **50-64 yrs** | **65-74 yrs** | **75+ yrs** |  |
| **Indirect costs** | Workforce participation rate | 0.0% | 60.7% | 76.6% | 64.8% | 24.7% | 8.3% | [32] |
|  | Labor cost per week (USD) | 0 | $750 | $1,072 | $1,131 | $989 | $989 | [33] |
|  | **Working time lost (days) among patients in:** | | | | | | | |
|  | Outpatient care | 10.0 | | | | | | [34] |
|  | General ward without IMV | 20.1 | | | | | | [19, 35] |
|  | General ward with IMV | 183 | | | | | | [36, 37] |
|  | ICU without IMV | 23.6 | | | | | | [19, 35] |
|  | ICU with IMV | 183 | | | | | | [36, 37] |
|  | Patients with PASC | 60.0 | | | | | | *Assumption* |
|  | **Productivity loss (USD) among patients in^b^:** | | | | | | | |
|  | Outpatient care | $0 | $650 | $1,173 | $1,047 | $348 | $117 |  |
|  | General ward without IMV | $0 | $1,307 | $2,359 | $2,104 | $700 | $236 |  |
|  | General ward with IMV | $0 | $11,899 | $21,473 | $19,153 | $6,377 | $2,146 |  |
|  | ICU without IMV | $0 | $1,535 | $2,769 | $2,470 | $822 | $277 |  |
|  | ICU with IMV | $0 | $11,899 | $21,473 | $19,153 | $6,377 | $2,146 |  |

COVID19, coronavirus disease 2019; ICU, intensive care unit; IMV, invasive mechanical ventilation; PASC, post-acute sequelae of COVID-19; USD, United States dollars

^a^ Participation rate and labor cost were defined none for the age group of 5-17 yrs including 5-11 for scenario analysis.

^b^ Productivity loss (USD) among susceptible and protected patients were same.

2.9. **Health utilities**

Disutility weights derived from the patient data of other infectious diseases such as influenza and *Clostridium difficile*, were applied to individuals experiencing COVID-19 symptoms and sequelae after getting infected. Whereas age-specific utility values for individuals without infection were used from Sullivan et al., which provided community-based EuroQoL-5-dimension (EQ-5D) scores. As no general population utility norms were available for the 12-17 age group, weights for the 18-29 age group were adopted. These disutility weights were applied to the duration of COVID-19 specific symptoms and sequelae for individuals in the susceptible and protected (vaccinated or recovered) health states. The durations of COVID-19 related outpatient and inpatient treatment, were sourced from the Premier Healthcare database analysis by Di Fusco et al. We assigned the same duration for all age groups per outcome of interest as Di Fusco results were not age specific.

A short-term QALY impact was included for all individuals receiving mechanical ventilation to reflect the longer recovery time and impact on quality of life. Similarly, QALY loss associated with developing post-acute sequelae of COVID-19 (PASC) was also included. Long-term QALY loss was estimated using the relative ratio of published disability weights for post-acute consequences to moderate community cases, multiplied by the assumed QALYs lost per symptomatic case. Long-term QALY loss associated with early death was included on a lifetime horizon.

**Table S14: Health utility inputs**

| **Category** | **Input description** | **Base Case** | **Source** |
| --- | --- | --- | --- |
| **Utility input** | **Utility weights for the age groups** |  |  |
|  | 5-11^a^ | 0.92 | *Assumed same as of age 18-29* |
|  | 12-17 | 0.92 |  |
|  | 18-29 | 0.92 | [38] |
|  | 30-49 | 0.89 |  |
|  | 50-64 | 0.84 |  |
|  | 65-74 | 0.81 |  |
|  | 75+ | 0.86 |  |
|  | **COVID-19-related disutility** |  |  |
|  | COVID-19 symptoms | -0.19 | [39] |
|  | General ward hospitalization without IMV | -0.3 | [40] |
|  | General ward hospitalization with IMV | -0.6 | *Assumption (same as ICU stay with IMV)* |
|  | ICU stay without IMV | -0.5 | [40] |
|  | ICU stay with IMV | -0.6 |  |
|  | **Duration of decreased utility (days)** |  |  |
|  | Duration of COVID-19 symptoms for outpatients | 5.0 | *Assumption based on the WHO-China Joint Mission on COVID-19*. [35] |
|  | **Duration of COVID-19 inpatient stay** |  |  |
|  | General ward (without/with IMV) | 5-11: 3.1/2.9  12+: 6.1/12.1 | 5-11: [19]  12+: [19] |
|  | ICU (without/with ventilation) | 5-11: 5.5 / 21.5  12+: 9.6/18.6 |  |
|  | **QALY Decrements for Outpatient and Ventilation** |  |  |
|  | Long term QALY effect of IMV | -0.06 | *Assumption based on Hamel et al.* [41] |
|  | QALY effect of PASC | -0.034 | [42] |
|  | QALY effect of post-acute consequences | -0.219 | [43] |
|  | QALY effect of moderate community cases | -0.051 |  |
|  | QALY effect of symptomatic cases | -0.008 | [44] |
|  | COVID-19 symptoms | -0.003 | [44] |

COVID-19, coronavirus disease 2019; ICU, intensive care unit; IMV, invasive mechanical ventilation; PASC, post-acute sequelae of COVID-19;

QALY, quality-adjusted life year. ^a^ Scenario analysis in the age group of 5-11.
